# Conventional and Bayesian workflows for clinical prediction modelling of severe Covid-19 outcomes based on clinical biomarker test results: LabMarCS: Laboratory Markers of COVID-19 Severity - Bristol Cohort

**DOI:** 10.1101/2022.09.16.22279985

**Authors:** Brian Sullivan, Edward Barker, Philip Williams, Louis MacGregor, Ranjeet Bhamber, Matt Thomas, Stefan Gurney, Catherine Hyams, Alastair Whiteway, Jennifer A Cooper, Chris McWilliams, Katy Turner, Andrew W. Dowsey, Mahableshwar Albur

## Abstract

We describe several regression models to predict severe outcomes in COVID-19 and challenges present in complex observational medical data. We demonstrate best practices for data curation, cross-validated statistical modelling, and variable selection emphasizing recent Bayesian methods. The study follows a retrospective observational cohort design using multicentre records across National Health Service (NHS) trusts in southwest England, UK. Participants included hospitalised adult patients positive for SARS-CoV 2 during March to October 2020, totalling 843 patients (mean age 71, 45% female, 32% died or needed ICU stay), split into training (n=590) and validation groups (n=253). Models were fit to predict severe outcomes (ICU admission or death within 28-days of admission to hospital for COVID-19, or a positive PCR result if already admitted) using demographic data and initial results from 30 biomarker tests collected within 3 days of admission or testing positive if already admitted. Cross-validation results showed standard logistic regression had an internal validation median AUC of 0.74 (95% Interval [0.62,0.83]), and external validation AUC of 0.68 [0.61, 0.71]; a Bayesian logistic regression (with horseshoe prior) internal AUC of 0.79 [0.71, 0.87], and external AUC of 0.70 [0.68, 0.71]. Variable selection performed using Bayesian predictive projection determined a four variable model using Age, Urea, Prothrombin time and Neutrophil-Lymphocyte ratio, with a median internal AUC of 0.79 [0.78, 0.80], and external AUC of 0.67 [0.65, 0.69]. We illustrate best-practices protocol for conventional and Bayesian prediction modelling on complex clinical data and reiterate the predictive value of previously identified biomarkers for COVID-19 severity assessment.

## Introduction

Globally as of 26 April 2023, there have been 764 million confirmed cases of COVID-19, including 6.9 million deaths, with 24.6 million cases in the UK, resulting in over 207,000 deaths (WHO Coronavirus (COVID-19) Dashboard, https://covid19.who.int/). COVID-19 has a wide spectrum of clinical features ranging from asymptomatic to severe systemic illness with a significant attributable mortality, while clinical manifestations are variable especially in the most vulnerable groups and immunocompromised people [1]. COVID-19 is a multi-system disease resulting in the derangements of homeostasis affecting pulmonary, cardiovascular, coagulation, haematological, oxygenation, hepatic, renal and fluid balance [2, 3, 4, 5, 6, 7, 8, 9, 10, 11, 12, 13, 14, 15, 16]. Although the majority of people with COVID-19 will have mild or no symptoms, a small but significant proportion will suffer from a severe infection needing hospitalisation for supportive care, oxygen, or admission to intensive care units (ICU) for respiratory support.

Early identification of hospitalised COVID-19 patients who are likely to deteriorate, i.e. transfer to ICU or who may die, is vital for clinical decision making. Healthcare systems across the world including highly developed countries continue to face challenges in terms of capacity and resources to manage this pandemic, as lock down measures have been relaxed, including opening of schools and businesses.

To date, published prediction models have evaluated case-level factors that might predict poor outcomes (critical illness or death). A recent living systematic review [17] identified 265 prognostic models for mortality and 84 for progression to severe or critical state. The majority of the studies looked at vital signs, age, comorbidities, and radiological features. Models were unlikely to include a broad range of variables concerning co-infection, biochemical factors (outside of C-reactive protein), and other haematological factors on an individual patient level. Most of the prognostic models did not describe the target population or care setting adequately, did not fully describe the regression equation, showed high or unclear risk of bias and/or were inadequately evaluated for performance.

### Goals

The present study analyzes a range of laboratory blood marker values across metabolic pathways affected by COVID-19 infection (i.e. a core set of biomarkers feasible for clinical collection) and evaluates predictive models of severe outcomes. The main objectives of the study are: (1) Examine statistical associations of routinely measured physiological and blood biomarkers, and age and gender, to predict severe COVID-19 outcomes. (2) Develop cross-validated logistic regression prediction models using the candidate biomarkers, highlighting biomarkers worthy of future research. (3) Use variable selection techniques including least absolute shrinkage and selection operator (LASSO) regularisation [18] and Bayesian Projective Prediction [19] to illustrate the process of creating a reduced model that maintains reasonable performance and is more feasible to use clinically. (4) In each of these steps, demonstrate best analytic practices for explaining clinical data curation and statistical modelling decisions, with an emphasis on showcasing the capabilities of recent Bayesian methods.

## Methods

### Study Cohort and Demographics

Pseudonymised data was obtained from Laboratory Information Management Systems (LIMS) linking patient data for laboratory markers to key clinical outcomes. Three hospitals in the Southwest region of England, UK, participated in the study, two of which were tertiary teaching hospitals and the third was a district general hospital (DGH). A system-wide data search was conducted on the LIMS for all patients who tested positive for SARS-CoV-2 by polymerase chain reaction (PCR) at these three hospitals during the first wave of COVID-19 pandemic (01/03/2020 to 31/10/2020). The serial pathology data collected as a part of standard of care of patients admitted with/for COVID-19 were included-bacteriology, virology, mycology, haematology, and biochemistry. All patients testing negative for SARS CoV-2 by PCR were excluded. All laboratory markers including clinical outcomes from LIMS were extracted and the final dataset was anonymized with no patient identifying data to link back.

### Inclusion and exclusion criteria

We included all adult patients admitted to study hospitals and tested positive for SARS CoV-2 by PCR. Pediatric patients (<18 years old) and staff/healthcare workers and their house-hold contacts were excluded. Figure 1 depicts the decision flow for inclusion and exclusion of patient data.

**Figure 1:**
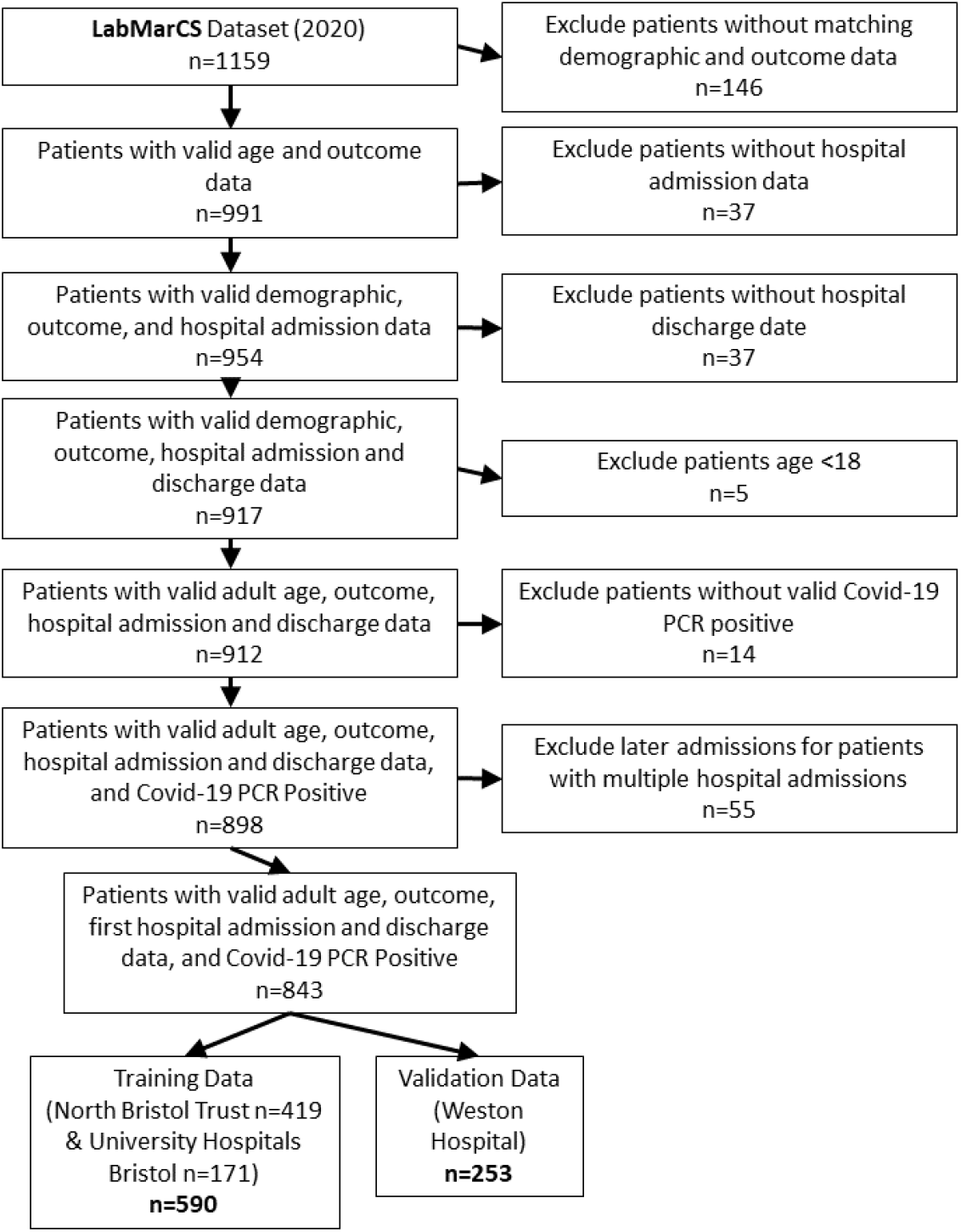
Flowchart of patient exclusion and inclusion criteria. The initial set of 1159 candidate patients was narrowed to a training set (n=590) and a validation set (n=253).

### Data Covariates

The LabMarCS dataset includes a variety of host, clinical severity indices, microbiological, immunological, haematological and biochemistry parameters used as predictive variables in the regression models. A full list of recorded data items is shown in Figure 2

**Figure 2:**
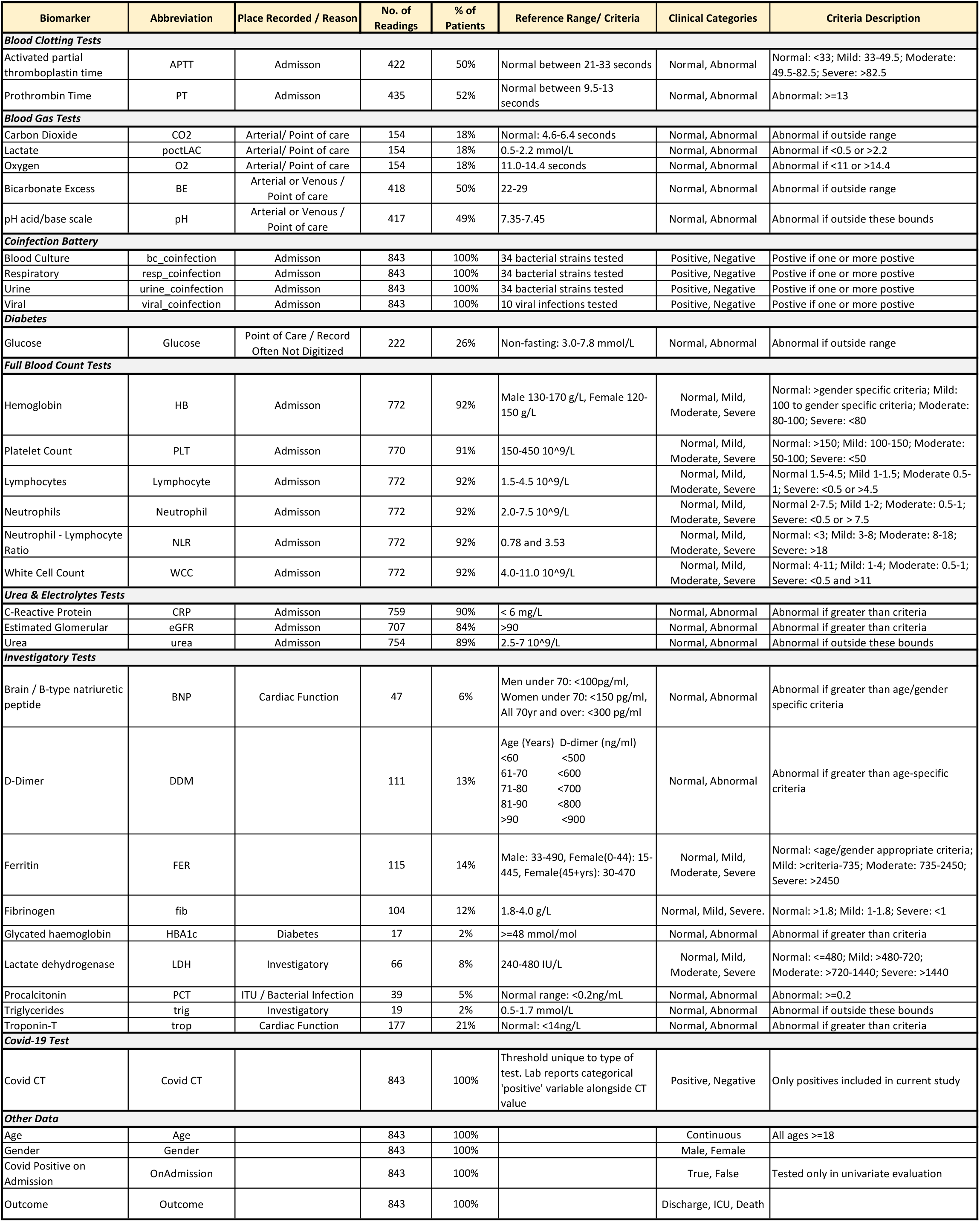
Variables recorded in the LabMarCS dataset, including plain text description, abbreviation, place of record, frequency in the dataset, and criteria used for converting continuous readings into categorical values.

### Outcomes

For all sites, the primary prediction outcome was death or transfer to the ICU within 28 days of the critical date. This critical date was either the point of admission to hospital, or the date of the first positive COVID-19 PCR test result if the patient was already admitted. This generally corresponds to WHO-COVID-19 Outcomes Scale Score 6–10 (severe) versus 0–5 (mild/moderate) [20].

### Patient Timelines

The collected laboratory biomarkers are continuous measures and provide a time-series representation of the course of a patient’s admission. Figure 3 shows an example of a single patient’s readings over the course of 18 days between testing positive for COVID-19 and being released from hospital care. This provides a representative example of the heterogeneity seen in our dataset, i.e. not all tests are taken and others are taken regularly or intermittently (further examples in Supplementary Figures S2 - S6).

**Figure 3:**
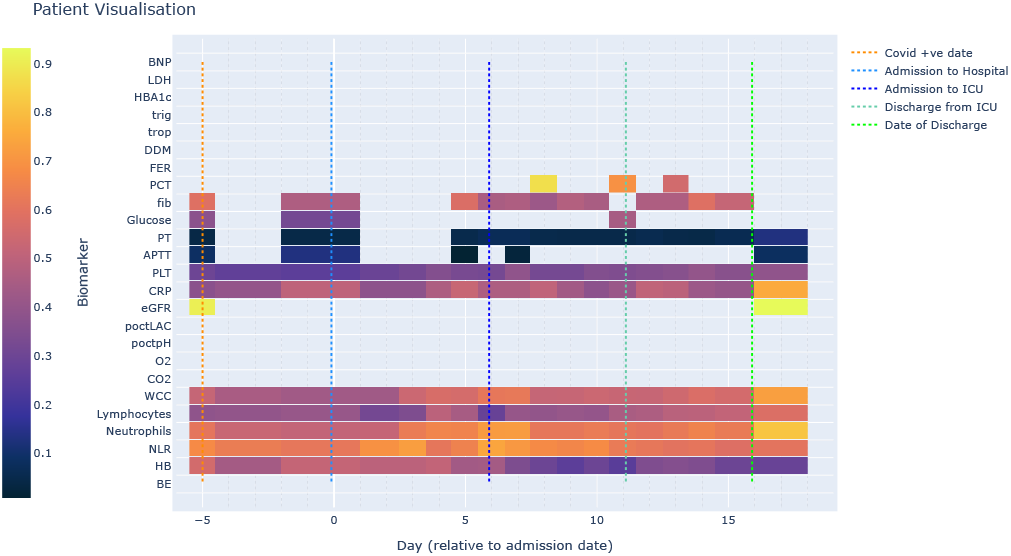
Example a single patient’s time series laboratory biomarker data. See Figure 2 for biomarker abbreviations. Biomarker results are normalised to span 0 to 1 via offsetting by the absolute value of the minimum value and dividing by the maximum value.

### Transformation of Biomarker Data

Prediction modelling of irregularly sampled time-series data is a challenging open research question [21]. In this study we focused on established and available tools for conventional and Bayesian prediction. To balance inclusion of test data not available on the day of admission and the need for clinical decisions to be guided soon after admission, we chose to consider the first value recorded for each biomarker within three days of their ‘critical date’. We additionally considered the worst or best readings within 1, 5 or 7 days, but found the first reading within 3 days to be the most realistic compromise. In addition, we transformed continuous biomarkers into categorical variables via reference ranges for clinical use in the typical healthy population ranges, see Figure 2. As an example, Figure 4 shows the histogram of readings for all values recorded for Neutrophils, including clinical thresholds to transform into categorical data. No missing data imputation was performed, instead missingness was coded as as an additional category ‘Test not taken’.

**Figure 4:**
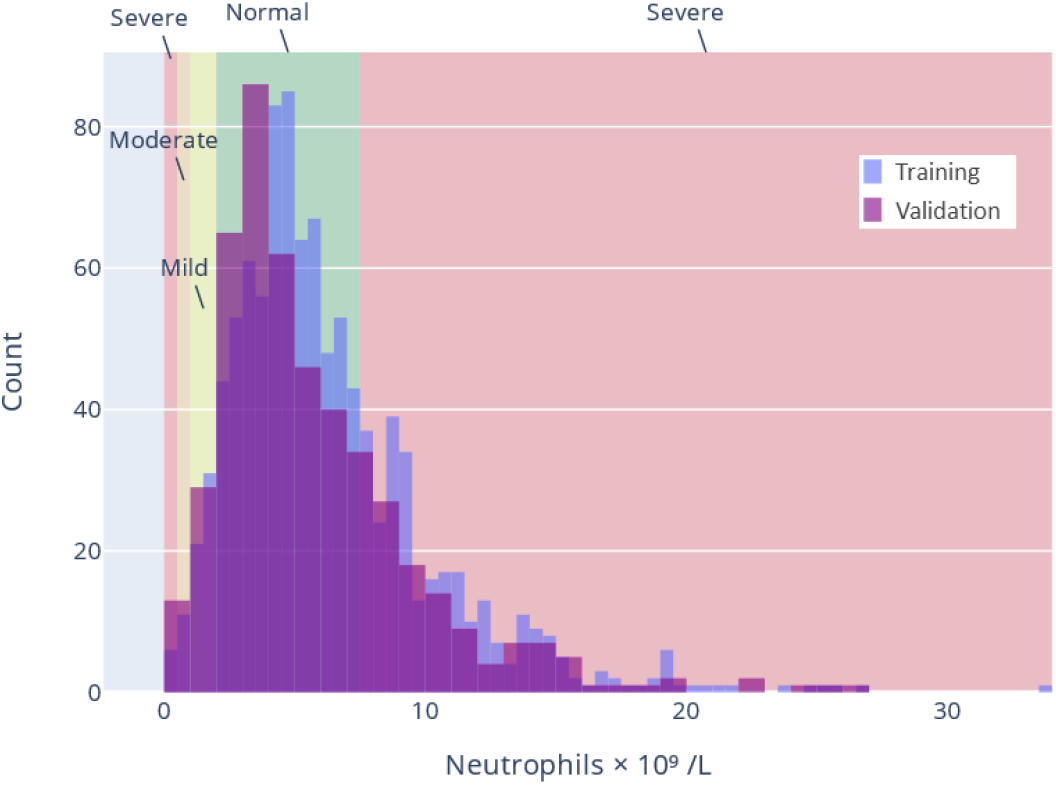
Example distribution of biomarker readings for Neutrophil Training and Validation Data. Vertical lines indicate clinical thresholds for bounds on Normal, Mild, Moderate, and Severe categorization.

For further elaboration these modelling choices and the challenges, please see Discussion section.

## Statistical Analysis

Analytics were carried out using the R statistical language (v4.13) and R Studio (Prairie Trillium release). We used the following packages: Standard logistic regression analyses used the R Stats GLM package (v3.6.2); LASSO analyses, GLMnet (v4.1-4); and for Bayesian analyses, BRMS (v2.17) and ProjPred (v2.1.2). Source code for this analysis pipeline can be found at https://github.com/biospi/LABMARCS.

### Analysis of Individual Biomarkers

Before running full regression models, we examined the independent contribution of individual biomarkers in predicting ICU entry or death via standard logistic regressions and Bayesian logistic regressions with either a flat (aka uniform) or horseshoe prior. This allowed calculation of p-values and odds ratios for each biomarker. A 5-fold cross-validation repeated 20 times was run for each biomarker to estimate median AUC and 95% interquartile intervals. Stratified cross-validation data shuffling was precomputed so all models used the same starting data. Stratified cross-validation separates patients by outcome (two groups of patients with severe outcomes and those without), and shuffles both into 5 groups (yielding an 80/20 training/test for each fold). These groups are combined ensuring all training and test datasets reflect the actual portion of patients with severe outcomes for that particular biomarker and not a random sample of that portion, which helps guarantee model convergence for biomarkers with high data missingness.

Only complete cases of training data available for each biomarker were considered, i.e. we did not include data for variables marked ‘Test not taken’, to focus on the predictive power of test results. Each individual biomarker model included age and gender (except univariate age and gender models) and were compared against a standard model including only age and gender. Regressions were fit using all associated dummy variables for a given biomarker (e.g. ‘Mild’, ‘Moderate’, ‘Severe’) using ‘Normal’ as the reference.

### Analysis Using All Valid Biomarker Data

After individual biomarker evaluation, logistic regression models considering all valid biomarkers (Prediction Using Individual Variables section) and demographic variables were fit to the data. Their predictions were tested via internal and external validation using the stratified cross-validation procedures detailed above, expect models were fit using all available training data using ‘Test Not Taken’ for absent data. The models include a standard logistic regression, a logistic regression regularised with LASSO, and two Bayesian models using a flat and a horseshoe prior [22]. LASSO encourages models to converge on sparse solutions with most coefficients set to zero to achieve variable reduction as discussed in the Reduced Variable Models section. Bayesian horseshoe prior models similarly encourage sparse solutions but without making hard decisions on variable inclusion - this can subsequently be performed using Projective Prediction.

### Analysis Using Reduced Variable Models

While a model using all biomarker data may have strong predictive power, it is clinically desirable to have a strong prediction with the least amount of biomarkers possible to save on resources devoted to biomarker collection. We used two methodologies to choose reduced variable models to predict COVID-19 severe outcomes, LASSO and Bayesian Projective Prediction.

LASSO is an optimization constraint that shrinks parameters according to their variance, reduces over-fitting, and enables variable selection [18]. The optimal degree of regularisation is determined by tuning parameter *λ* within each cross-validation fold through a nested cross-validation step. LASSO has a drawback of having biased coefficient and log-odds estimates, as such after evaluating LASSO models we run a standard GLM model on the reduced biomarker panel selected with the LASSO.

To evaluate LASSO coefficient estimates, we performed repeated nested stratified cross-validation (5-folds the for the inner LASSO loop; 5-folds for the outer loop, and 20 repeats). For a particular dataset fit, LASSO optimises for a sparse representation with many coefficients set to zero. Across cross-validated trials these variables will vary. LASSO fits are statistically biased and are better suited as a guide for variable selection in a reduced variable standard GLM. As recommended in Heinze et al [23], we consider the frequency of how often a particular biomarker has a coefficient greater than zero and count across cross-validation trials.

For determining unbiased effect sizes for the reduced variable set with a standard GLM, it was chosen that if at least one categorical level for a particular biomarker (e.g. ‘Severe’) was selected by the LASSO, all levels for that biomarker were included in the model. This resulted in a final set of ‘LASSO inspired’ variables that were then fit with standard logistic GLM. Note this approach, and more generally fitting multiple models to the same dataset, is subject to the problem of selective inference (aka multiple comparison error), see [24, 25] and the related R package [26]. Given our focus is not on reporting p-values, but instead cross-validation and generalisation from training data to validation data, these concerns are minimized.

The second variable selection method explored was Bayesian Projective Prediction [19], a technique for assessing reduced variable models against a complete ‘reference’ model, which in our case is a Bayesian logistic regression with a horseshoe prior [22]. Priors such as the horseshoe can be applied to provide adaptive shrinkage to covariates in Bayesian models directly so that full posterior distributions of odds estimates can be generated in an unbiased way. Unlike the LASSO, this does not shrink coefficients to zero exactly as the inherent uncertainty is not ignored. To perform hard variable selection, the recent approach of Projective Prediction can be used to compare the fit of submodels of the reference model through projections and approximate leave-one-out (LOO) cross-validation. Under the hood, Projective Prediction uses LASSO (or forward search) to select submodels for comparison, but retains the Bayesian inference for coefficient ranking and odds-ratio estimates. This approach allows one to evaluate the trade-off between AUC performance and the number of variables included in the model and use a reduced model projection at a desired AUC cutoff. Further projective prediction allows the flexibility to train one model on all valid available data, perform variable selection, and then use any projected sub-model with reduced variables to predict outcomes for novel data. Projective prediction models were evaluated using cross-validation procedures described in prior sections. Note, the analysis of the projective prediction model using all training data uses LOO for variable selection, which is computationally intensive. To speed variable selection computation during our cross-validation analysis, we used ‘naive’ variable selection, which only considers the training data from current fold as is, and does not perform any further internal cross-validation (the projective prediction package allows naive, k-fold, and LOO).

## Results

### Cohort Description

The initial cohort included 1159 patients which was narrowed down to 843 patients who met all inclusion criteria described above, see Figure 1. 57% of patients were hospitalised for COVID-19 and the remainder had nosocomial infection. For our statistical models, the training cohort (n=590) was defined as all adults admitted to hospital and testing positive for SARS-Cov-2 by PCR, or testing positive while already admitted between March and October 2020. For external validation, we held the DGH cohort (n=253) out of training. Figure 5 depicts the distribution of ages and genders in the training and validation data sets. Patients in the training set had a mean age of 70, were 44% female, and 29% had severe outcomes. The validation set had a mean age of 75, were 47% female, and 38% had a severe outcome.

**Figure 5:**
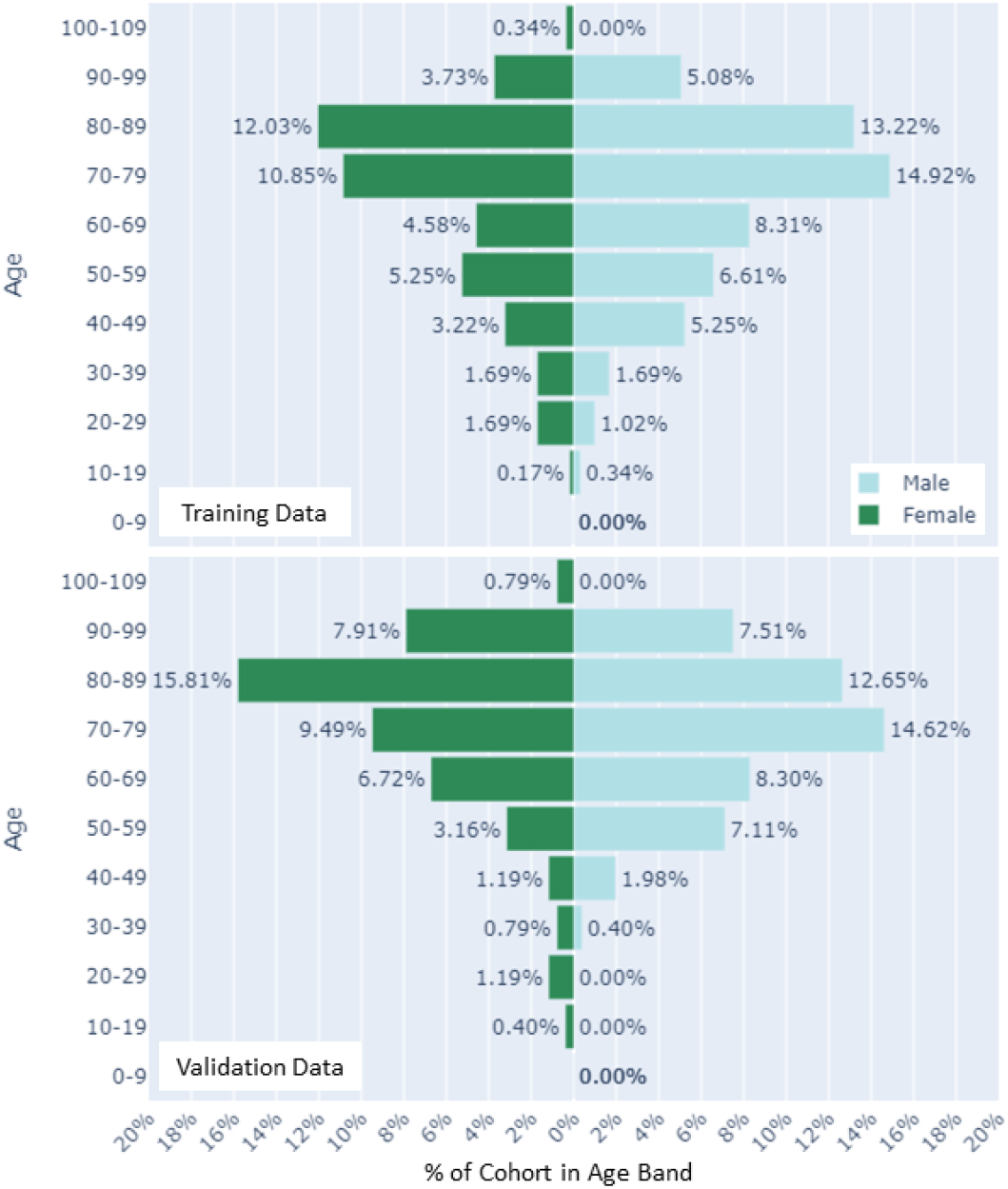
Distribution of age and gender for hospitalized patients with coronavirus disease 2019 (COVID-19) for (Top) training data (n=590) and (Bottom) hold out validation data (n=253) cohorts.

### Prediction Using Individual Variables

Figure 6 shows descriptive statistics on individual biomarker readings and their odds ratio contributions in a 5-fold 20-repeat stratified cross-validated logistic regression including the particular biomarker and age and gender. Figure 7 details performance using the area under the receiver operating characteristic curve (AUC) metric, comparing biomarker models (a particular biomarker plus age and gender) to a model using only age and gender. Due to the categorical representation of the biomarkers, individual levels may be significant while another is not (e.g. ‘Severe’ is a predictor, but ‘Mild’ is not). Statistically signifi-331 cant predictors (i.e. odds ratios deviating from one with p-value at 0.05 or lower) associated with increasing risk of a severe outcome (as shown in Figure 6) include age, and the biomarkers: Activated Partial Thromboplastin Time (Mild), Prothrombin time (Abnormal), blood pH (Abnormal), Haemoglobin (Severe), Platelet count (Moderate), Lymphocytes (Moderate, Severe), Neutrophils (Severe), Neutrophil-Lymphocyte Ratio (Mild, Moderate, Severe), C-Reactive Protein (Abnormal), Urea (Abnormal), and Troponin-T (Abnormal). Nosocomial transmission was included due to the high number of cases in our cohort but was not a significant predictor and excluded from further analyses. Due to small numbers preventing cross validation, Triglycerides, Glycated Haemoglobin, and Procalcitonin (also invalid due to being recorded only in ICU) were excluded from further analysis and require future research.

**Figure 6:**
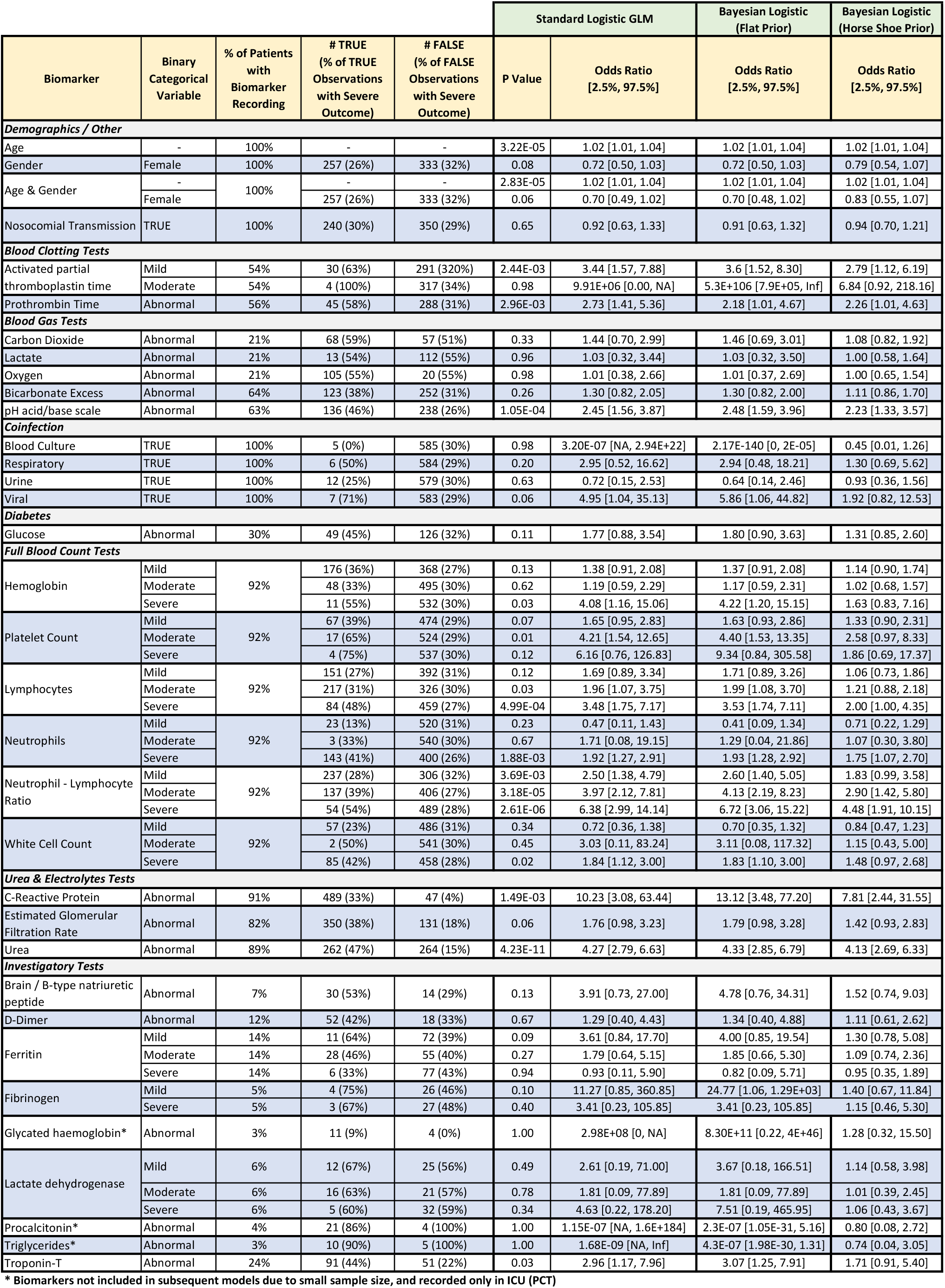
Individual biomarker evaluation including descriptive statistics and logistic regression model outcomes (Standard, Bayesian with flat prior, and Bayes with horseshoe prior), including age and gender (except univariate age and gender models). Regressions were fit using all associated dummy variables for a given biomarker (e.g. normal, mild, moderate, severe) and using only complete cases of training data, i.e. not using a variable for ‘Test not taken.’ Categorical variables use a reading of ‘Normal’ as a reference in the fitted model, except ‘Male’ used as the reference category for gender.

**Figure 7:**
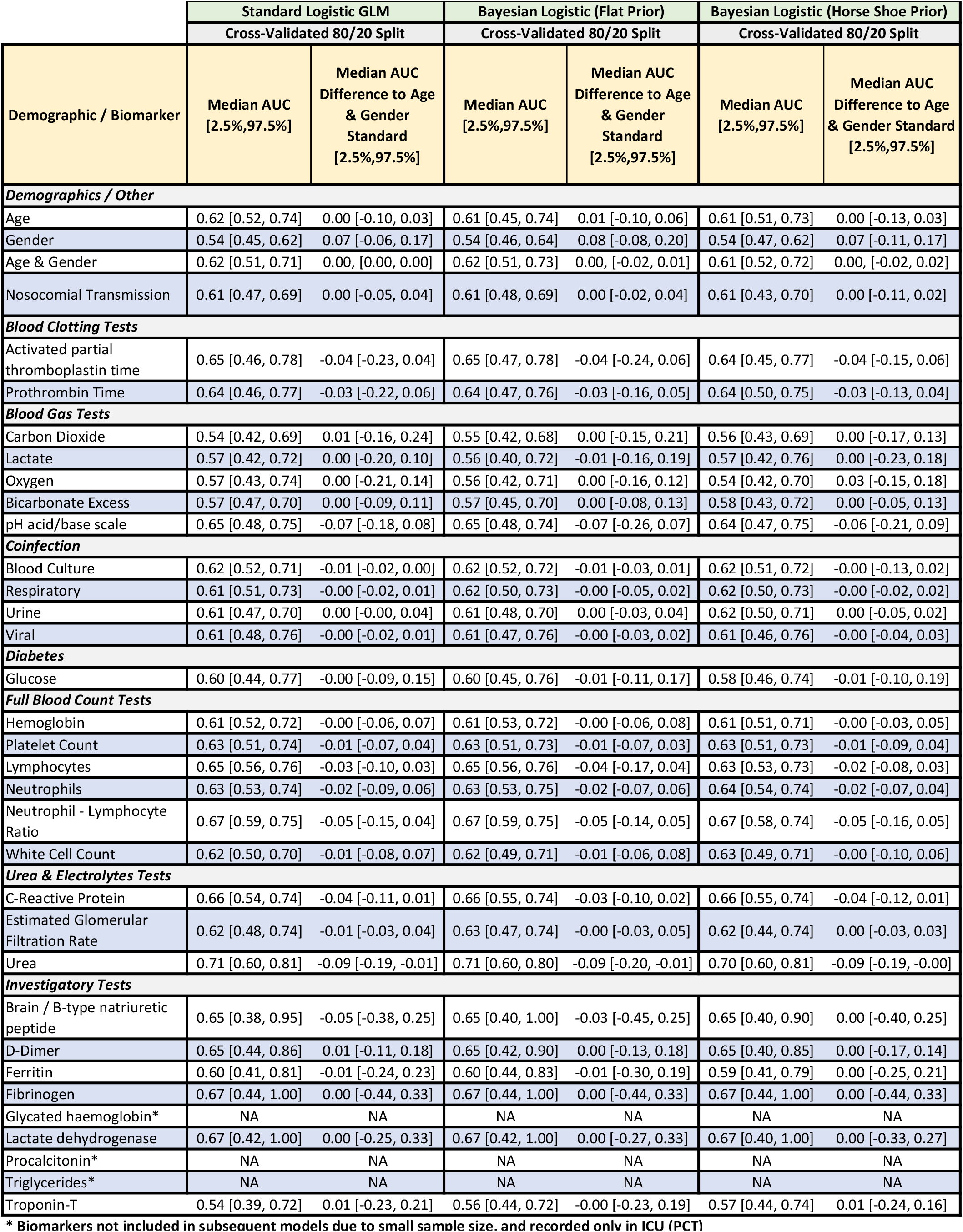
Predictive performance of the individual biomarker models in Figure 6 as described by the median area under the curve (AUC) in receiver operating curve (ROC) analysis and median difference between an Age and Gender reference model and the same model (negative values indicate the reference has worse performance) with the particular biomarker included (except univariate age and gender models). Regressions were fit using all associated dummy variables for a given biomarker (e.g. mild, moderate, severe) and using only complete cases of training data (n=590), i.e. not using a variable for ‘Test not taken.’ 95% interquantile ranges calculated via 5-fold cross-validation with 20 repeats (100 models total). Categorical variables use a reading of ‘Normal’ as a reference in the fitted model, except ‘Male’ used as the reference category for gender.

### Regression Models Using All Valid Biomarker Data

Each model was evaluated via 5-fold stratified cross-validation with 20 repeats (100 models total). As such, each model is trained with a randomised sample of 80% of the training data set (n=472). Internal validation evaluates model predictions on the 20% (n=118) held out. External validation uses the same model, but is instead tested on the never trained on validation data set (n=253). Missing data for each biomarker is coded as ‘Test Not Taken’ and is included as a predictor variable. Figure 8 shows the performance of these models (AUC, Sensitivity, Specificity). For comparison, Figure 9 shows the performance of each model using all valid training data (n=590) and testing on the same data (internal validation) and testing on the held out external validation data (n=253).

**Figure 8:**
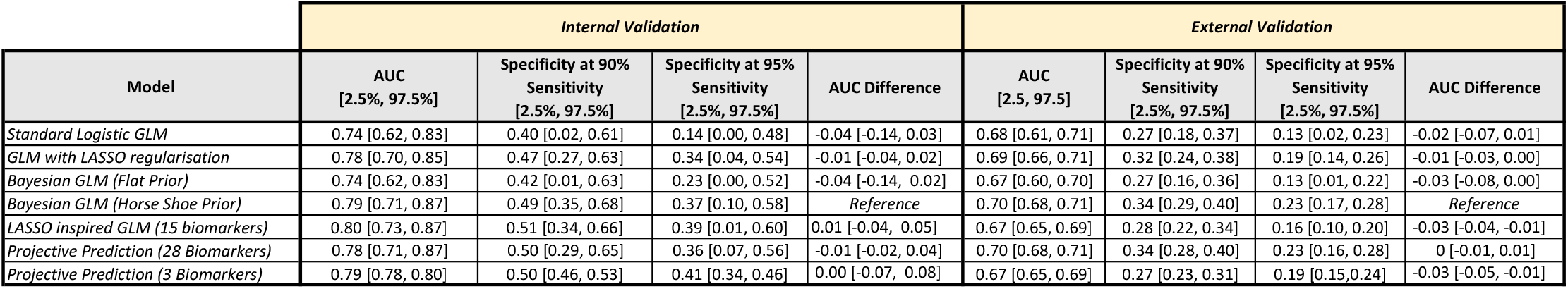
Cross-validated performance of models trained using valid biomarker data. 95% inter-quantile ranges are presented for each estimate. Specificity is obtained by evaluating at a set sensitivity of either 90% or 95%. All reduced variable models include age, and a stated number of biomarkers. The reduced variable standard GLM uses 15 biomarkers that had non-zero coefficients on >=50% LASSO Cross-validation trials. If at least one categorical level for a particular biomarker (e.g. severe) met this requirement, all levels for that biomarker were included in the model. The 3 biomarker projective prediction model uses all categorical levels for Urea, PT, and NLR. Pairwise AUC difference is presented in comparison to the Bayesian (Horse shoe prior) model.

**Figure 9:**
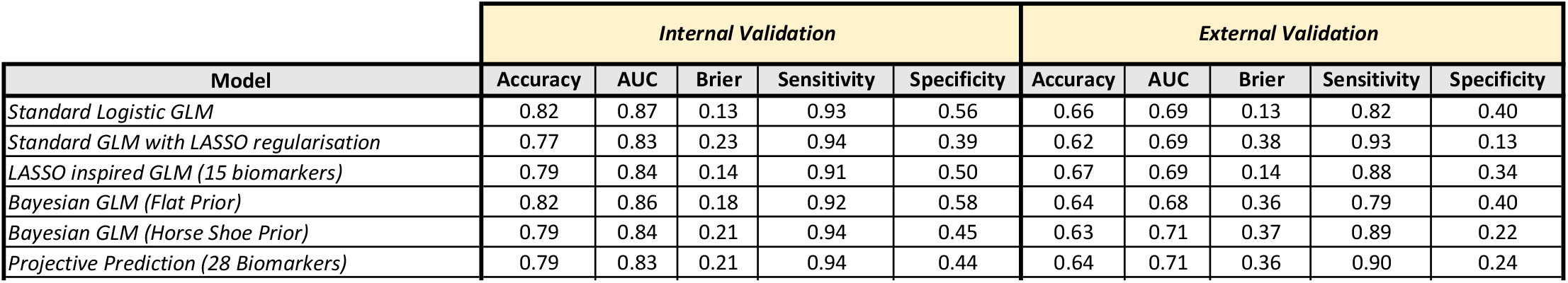
Performance of models using all valid biomarker data trained on all training data available (n=590). Internal validation is trained on all of the training data and tested on the same. External validation uses the same model and is tested on held out validation data set (n=253). Missing data for each biomarker is coded as ‘Test Not Taken’. Specificity and sensitivity evaluated using a probability threshold of 0.5 (i.e. assumes a well-calibrated model). All reduced variable models include age, and a stated number of biomarkers. The reduced variable standard GLM uses 15 biomarkers that had non-zero coefficients on >=50% LASSO Cross-validation trials. If at least one categorical level for a particular biomarker (e.g. severe) met this requirement, all levels for that biomarker were included in the model. The 3 biomarker projective prediction model uses uses all categorical levels for Urea, PT, and NLR.

Models trained on the full data have improved AUC scores, but do not provide a direct uncertainty estimate. For a single model this could be done via bootstrapping, but would not include uncertainty in model fit. Instead we compute interquantile ranges using 5-fold 20-repeat cross-validation of models. Cross-validation results provide 95% inter-quantile ranges that clearly illustrate that in general, all models perform similarly, with a median AUC in the mid 0.70’s in internal validation, and near the high 0.60’s in external validation. The Bayes model with horseshoe prior slightly outperforms all others, as shown in the AUC difference column showing the distribution of pair-wise differences across folds and repeats. The calibration of the models is reasonably good on the full data, all training data, but has poor calibration on the validation set, see Supplementary Figure S9.

### Reduced Variable Models

The models detailed above are moderately good predictors of severe COVID-19 outcomes, but for clinicians with limited time and resources, reduced models can balance predictive performance with ease of clinical use by using only the most informative biomarkers. To address this, we use two variable selection approaches, LASSO and projective prediction, that allow the creation of reduced models with fewer biomarkers but similar performance to the larger models.

### LASSO Models

After performing 5-fold 20 repeat cross-validation we examined the frequency of how often a particular biomarker has a coefficient greater than zero and count across cross-validation trials. Figure S10 shows the frequency of variables having a coefficient great than zero in the cross-validated LASSO analysis. If we select variables that appear at least 50% of the time, our reduced model would include: Age, CRP (abnormal), FER (mild), FIB (mild), HB (severe), PLT (mild, moderate, severe), Lymphocytes (Severe), Neutrophils (Mild, Severe), NLR (Severe), APTT (mild, moderate), PT (abnormal), blood pH (abnormal), Urea (abnormal), and positive viral and blood culture co-infections.

For the reduced variable standard GLM, this resulted in a model using the 15 biomarkers above for all categorical levels, and was evaluated via both cross-validation and as fit to all available training data. This model had performance very similar to the models using all valid biomarker data, with a median external validation AUC of 0.68 [0.63, 0.72], see Figures 8 and 9.

Note, ‘Test Not Taken’ is a significant predictor for LDH and Lactate on over 50% of cross-validation trials. The potential significance of missing data is complex and is addressed in the Discussion section. Due to this confounding, biomarkers whose top predictive contribution was from ‘Test Not Taken’ were excluded from both LASSO reduced variable models and projective prediction models described below.

### Projective Prediction Models

When all biomarkers were considered, projective prediction identifies the following predictors in the top 20, in order of contribution to AUC: Urea (abnormal), Age, PT (abnormal), NLR (Severe), pH (abnormal), Lymphocytes (severe), APPT(mild), eGFR (abnormal), Neutrophils (Severe), APPT(moderate), CRP (abnormal), DDM (abnormal), Hemoglobin (severe). Thus age and 12 biomarkers are candidates for a reduced model. Note, several predictors of ‘Test Not Taken’ were also selected including Lactate, O2, CO2, LDH, Ferritin and Fibrinogen. As mentioned above, these biomarkers are set aside due to this confounding. Supplementary Figures S11 and S12 display the output from projective prediction ranking the contribution of each variable to the model. A model using a projection incorporating all biomarker and demographic data is equivalent to the standard Bayesian GLM we evaluated in the prior section, see Figures 8 and 9.

Reduced variable projections were evaluated by manual inspection of AUC performance among groups of models using the top biomarkers. Guided by the projective prediction ranking, we ran a model using only the top biomarker, using only the top two, the top three, and so on. As described above we omit biomarkers with significant contributions from ‘Test Not Taken’ and include all categorical levels for a given biomarker as long as one level is highly ranked. Ultimately, we found a 3 biomarker projective prediction model using age and including urea, prothrombin time, neutrophil-lymphocyte ratios had similar performance to larger models with a median internal validation AUC of 0.79 [0.78, 0.80], and external validation AUC of 0.67 [0.65, 0.69], as shown in Figures 8 and 9. Odds ratios for the full Bayesian model and the reduced 3-biomarker model can be found in Supplementary Table S13. The calibration of the model is reasonably good on the training data but has poor calibration on the validation set, see Supplementary Figure S14.

## Discussion

### Challenges of Complex Medical Data

Curating the LabMarCS data is challenging as the data are heterogeneous in multiple ways. Biomarkers are recorded for different reasons, e.g. routine upon admission, investigatory tests, or tests primarily or exclusively taken in ICU. Further, some biomarkers are typically recorded together (but not always) as part of a test suite, including: Urea and electrolytes, full blood count, COVID-19 and co-infection swab test, blood clotting, and blood gas tests (arterial or venous). The schedule when some these markers are recorded vary by patient and clinical decision, leading to records being present in highly varying amounts, e.g. only 3% up to 100% of patients depending on the particular biomarker, see Supplementary Figure S1.

### Modelling Choices

When constructing and evaluating models, there are many choice points that should be explicitly highlighted with justification, be it based on convenience, computational complexity, clinical advice, or a heuristic. The space of potential models is vast and most studies will constrain the model search space, delineating why these choices are made will facilitate understanding and reproduction by other researchers. These include key choices relating to: patient inclusion/exclusion criteria, data missingness protocols, data transformations, training and validation data selection, and performance evaluation.

### Missing Data

Missingness, in the context of this study and in healthcare data more generally, can sometimes be informative and missing not at random (MNAR), with the presence or absence of a test correlated with the its measurement or the study outcome. Imputation of missing data relies on key statistical assumptions that imputed variables are missing at random (MAR) or missing completely at random (MCAR). Conversations with our clinical co-authors established some routinely collected biomarkers might be inferred to be MAR. However, the routines identified were specific to a small a subset of our cohort and not likely to extrapolate. We ultimately erred to be conservative and avoid all imputation, and instead include the presence/absence of missing values as a covariate itself [27, 28]. As such, in the current study we chose to use placeholders for ‘Test not taken’ if there was no recorded value for a particular biomarker within the evaluated 3-day window.

This approach allows the possibility that a ‘Test Not Taken’ may be a significant predictor. This has many potential meanings, as it may convey that when a patient is doing well and unlikely to experience a severe outcome, clinicians are unlikely to request some biomarker tests. Alternatively, if a patient is in palliative care and has a poor prognosis, a clinician may consider further testing unnecessary. As such, the likelihood of a test being administered may follow an inverted-U function as patients to healthy or too ill may not have tests administered. Furthermore, as our data was collected early in the pandemic, there may be other underlying clinical decisions or resource limitations that drove why some tests were taken but not others. Lastly, because we only consider results from the first 3 days from a patients critical date, it may be that some tests were simply taken later in a patient’s stay due to operational constraints, and hence may be more predictive as they were taken closer to the outcome. When these instances occurred, we were conservative and excluded biomarkers with ‘Test Not Taken’ as the most informative category from our reduced variable models.

### Data Transforms - Time Windows

Ideally clinicians make decisions based on readings on the day of admission. However, not all tests are administered on admission. To balance inclusion of test data not available on the day of admission and the need for clinical decisions to be guided soon after admission, we chose to consider the first value recorded for each biomarkers within three days of their ‘critical date’, i.e. date of admission if already COVID-19 positive, or if already in hospital, the date of testing COVID-19 positive. However, given the richness of the time series data collected, further research into models that leverage this extra information is needed.

Focusing on early detection reflects the intent for the model to improve early stage clinical decision making when potential treatments or changes in care may be introduced. This focus on the first reading in a 3-day interval loses information, but greatly simplifies the modelling approach. Note, this choice is not without risk of reducing statistical power, increasing the risk of false positives, and underestimation of the extent of variation in biomarker readings and outcomes between groups [29]. It is likely that representing biomarker data as time-series (assuming regular measures across patients) would add considerable information for continuous monitoring.

### Data Transforms - Continuous vs. Categorical

A key modelling decision must be made on whether to use continuous data or transformed categorical data. Clinicians often use biomarker thresholds to provide semantic categories (e.g. normal, mild, moderate, severe) which sometimes use non-linear or discontinuous mappings that require special care if using continuous data. While clinical thresholds are likely established with evidence, it may be the case that thresholds for one use may not apply to a novel use. This led [30, 31] to use machine learning approaches to build categorisation models on continuous biomarker data dependent on the training data at hand. However, using machine learning to establish categorisation thresholds on our biomarker data is difficult with a small training data set and the heterogeneity of biomarker recordings. If missing data imputation is performed, it raises another decision point on whether to impute the continuous or the transformed categorical data.

Another important factor to recognise is that some biomarkers lack a linear relationship between a reading and a semantic category. Biomarkers can have a lower and upper bound for what is considered normal, and both below and above this range reflects clinically meaningful yet sometimes separate abnormalities. The modelling needs to factor in non-linearity when persevering continuous data or trying to map to a categorical space. In our position, categorical transformation had an advantage, as they allowed us to collaborate with ICU consultants while using pre-established clinically acceptable ranges to define our categorisation, see Figure 2.

### Training and Validation Data Selection

There are multiple ways that our data set could be split between training and validation sets, e.g. randomly sampling 1/3 of the data to hold out as a validation set. Random selection of training data should in principle generate data more representative of the validation set left out. However, hospitals may have differing practices and non-stratified randomization may inflate performance at the cost of real world generalisation. We chose to separate our training and validation datasets by hospital to provide a stronger test of generalisation that should mimic generalisation to novel hospitals completely outside the original training data.

### Model Performance Evaluation and Dissemination

There are a variety of ways statistical model performance can be evaluated. Here we have chose here to emphasize cross-validated estimates of AUC, sensitivity, and specificity. Interquartile intervals over these measures reveal that the variety of models perform in similar ways. With a larger data set, tradeoffs may become more apparent. Model calibration on the validation set is a clear weak point. While the models have a reasonable calibration for training data, generalisation performance is weak and suggestive of the lack of sufficient data.

### Comparison to Contemporary Models

We found several biomarkers previously highlighted by other groups to have significant predictive power, including: Urea, Neutrophil-Lymphocyte Ratio, Lymphoctyes, APTT, eGFR, and CRP. Our highly reduced 3-biomarker model (plus age) uses Urea (highlighted by all prior models), Neutrophil-Lymphocyte ratio (highlighted by [32, 11, 31]), and APPT (highlighted by [31]). With a larger dataset, further vetting of these and other biomarkers would be possible, but it gives reassurance that despite limitations, we find similar predictive biomarkers.

### Advantages of Bayesian Modelling

While the predictive performance across models presented here is generally quite similar, the Bayesian horse shoe model had slightly better cross-validated predictive performance and there are several reasons for researchers to favor Bayesian approaches. Coefficients estimated via Bayes should on average deliver better predictive performance than standard GLM. Additionally, if a sparse model is needed, a horseshoe prior can provide advantages similar to LASSO without biased coefficient estimates. Computationally, Bayesian techniques can be slow due Markov Chain Monte Carlo used to sample the coefficient space. If one is interested in variable selection, projective prediction offers the ability to take a single Bayesian model fit, run a variable selection algorithm to rank variable contributions, and then arbitrarily create sub-model projections with any number of original variables. While the initial model fit and variable selection are computationally intensive, sub-model projections are fast to create and performance test.

## Summary & Conclusions

### Limitations

This is a retrospective cohort study in South-west England where case numbers have varied widely, and were below national incidence rates during the first wave. This results in less precise parameter estimates for prediction models (less power/smaller sample size) and likely reduced generalizability of the model to other settings. The timing of biomarker collection was highly varied both within and between patients, with many types of readings missing.

### Strengths

The primary strength of our study is the granularity of serial laboratory data available linked to clinical outcomes. This study was performed during the first wave where there was the original Wuhan strain circulating amongst the unvaccinated naïve population without any specific immunomodulating therapies such as steroids or antiviral agents, reflecting the “true” homeostasis derangements at a population level.

In particular, this study describes the variety of challenges present in complex medical data sets and how statistical bestpractices can be applied to such data, highlighting the benefits of recent Bayesian methodology. Our study reiterates the predictive value of previously identified biomarkers for COVID-19 severity assessment (e.g. age, urea, prothrombin time, and neutrophil-lymphocyte ratio). Both the full and reduced variable models have moderately good training performance, but improved external validation is needed for all models to be clinically viable. The methods presented here should generalise well to a larger dataset.

## Data Availability

Due to NHS data governance, all data produced in the present study are unavailable directly through the authors, but reasonable requests for data to Southwest England NHS can be arranged via the authors.

## Ethics approval

The study [IRAS project ID: 283439] underwent a rigorous ethical and regulatory approval process, and a favourable opinion was gained from Research Ethics Service, Wales REC 7, c/o Public Health Wales, Building 1, Jobswell Road, St David’s Park, SA31 3HB on 11/09/2020.

## Funding

This work is funded by Health Data Research UK via the Better Care Partnership Southwest (HDR CF0129), Medical Research Council Research Grant MR/T005408/1,the Elizabeth Blackwell Institute for Health Research, University of Bristol, and the Wellcome Trust Institutional Strategic Support Fund.

## Declaration of competing interest

The authors have no competing interests.

## Acknowledgements

This research was supported by the National Institute for Health and Care Research (NIHR) Applied Research Collaboration West (NIHR ARC West). The views expressed in this article are those of the author(s) and not necessarily those of the NIHR or the Department of Health and Social Care.

## Supplementary materials

**Figure S1:**
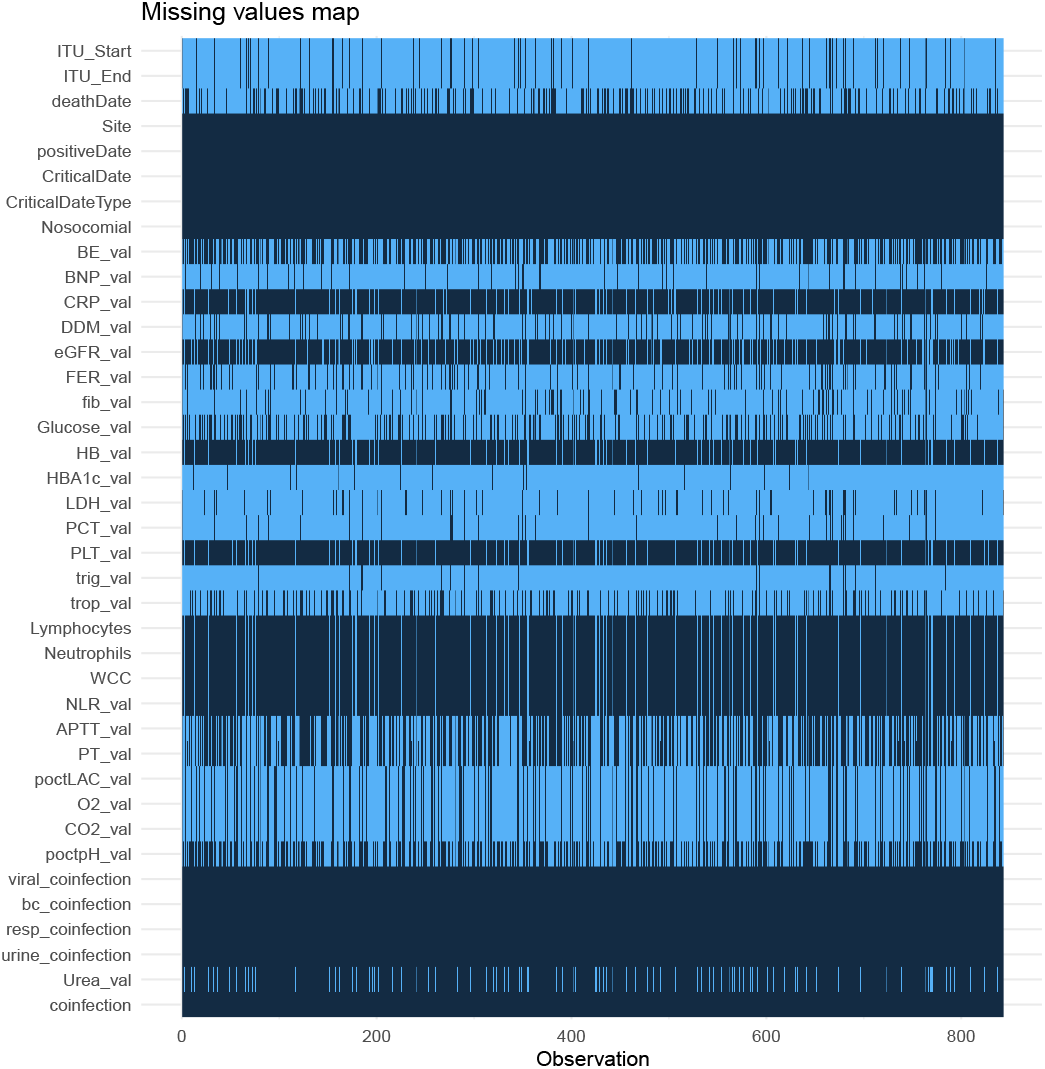
Heat map displaying missing values across recorded biomarkers. Light blue indicates a value is missing and dark blue indicate it is present

**Figure S2:**
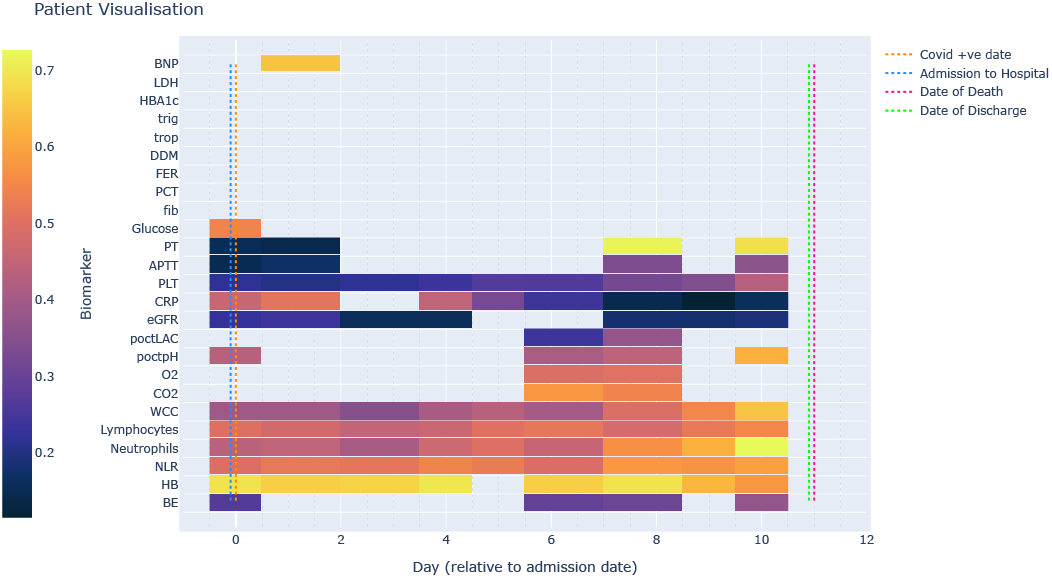
Example biomarker time series for a patient admitted to hospital COVID-19 positive and who subsequently died almost two weeks later.

**Figure S3:**
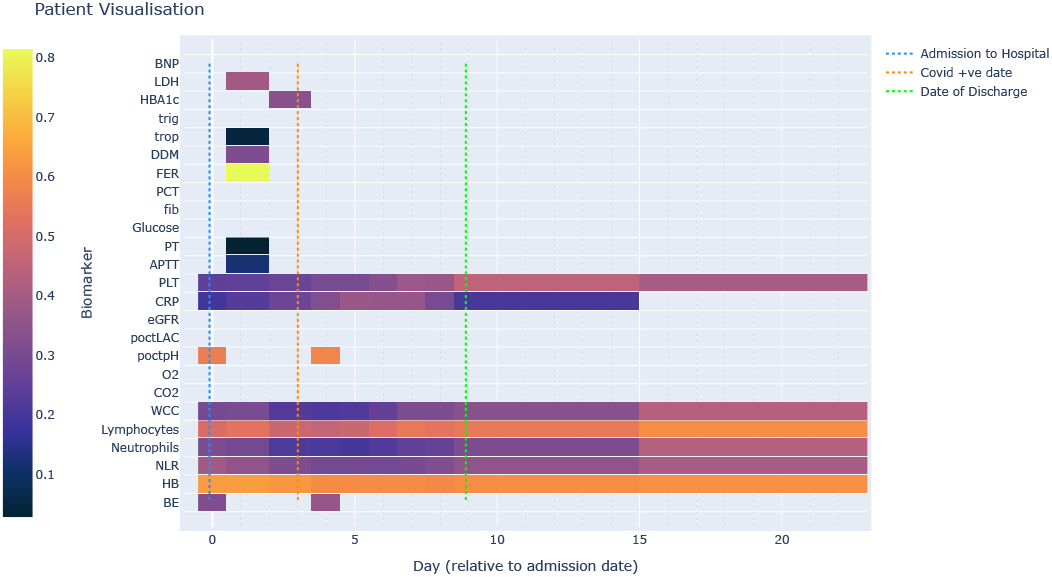
Example biomarker time series for a patient admitted to hospital with subsequent nosocomial transmission and discharge a week later.

**Figure S4:**
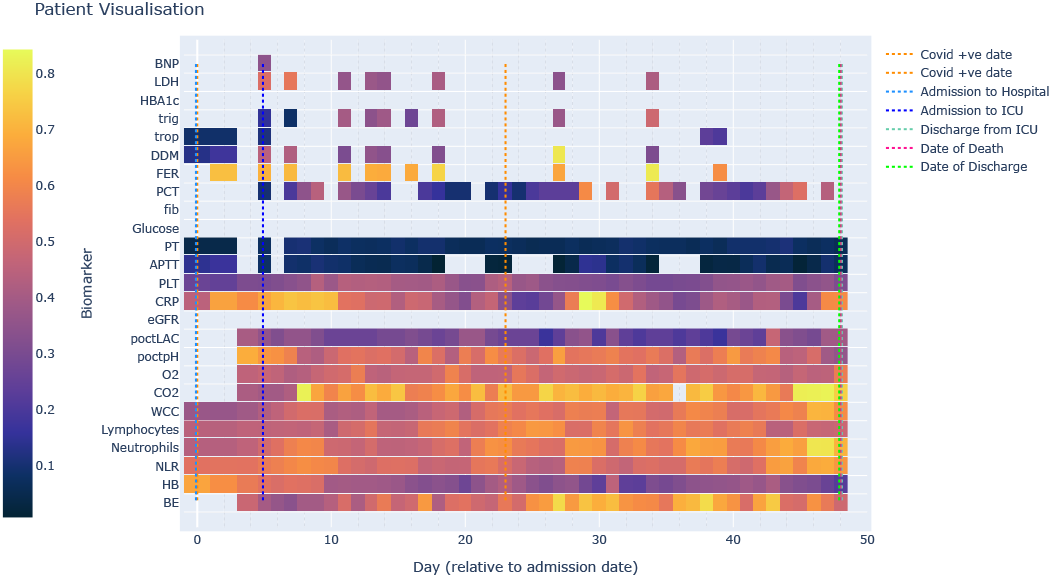
Example biomarker time series for a patient admitted to hospital COVID-19 positive, with subsequent entrance to ICU and death over one month later.

**Figure S5:**
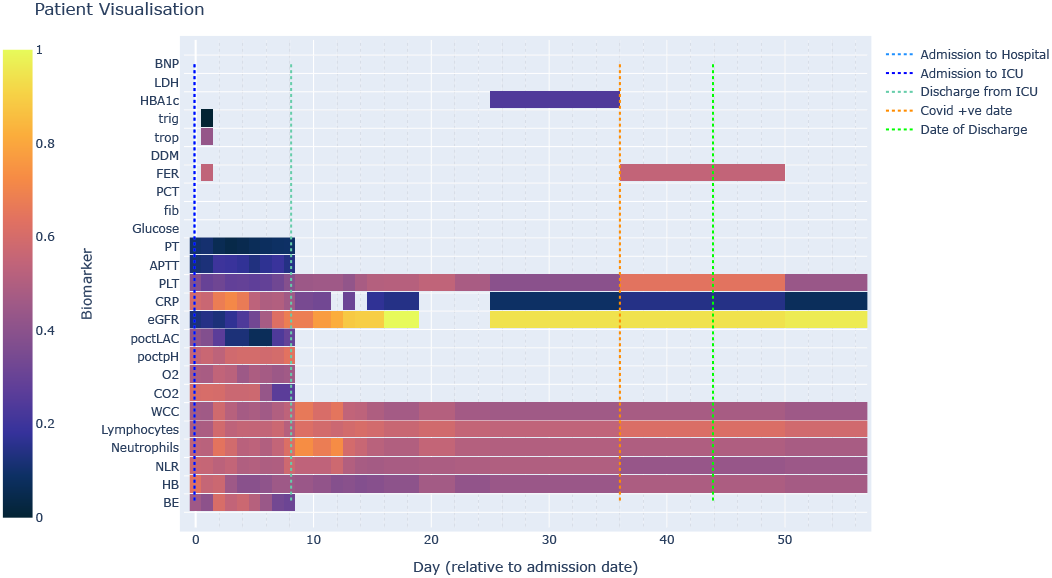
Example biomarker time series for a patient admitted to hospital and ICU, with subsequent nosocomial transmission and discharge about one week later.

**Figure S6:**
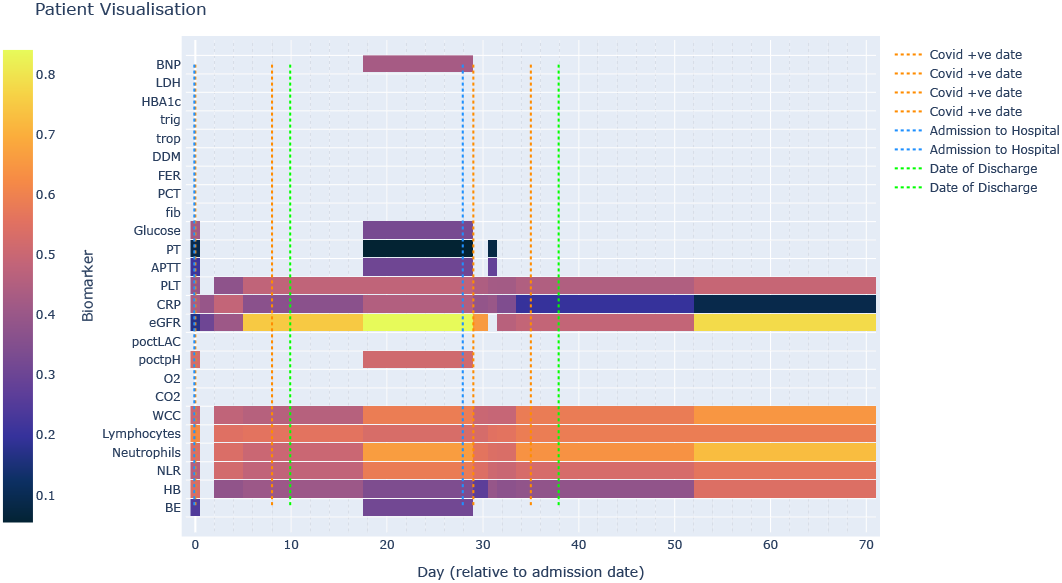
Example biomarker time series for a patient with two hospital admissions and testing COVID-19 positive on the first, with discharge almost two weeks after second admission.

**Figure S7:**
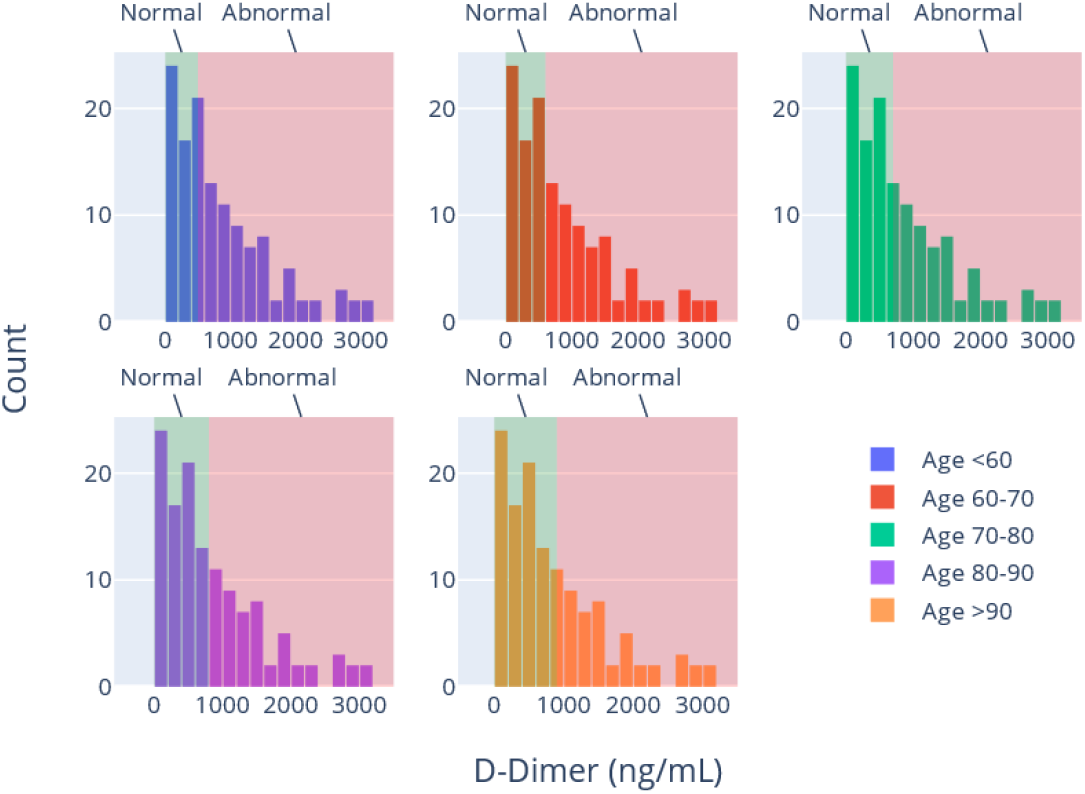
Distribution of D-Dimer readings with clinical classification requiring age and gender bands

**Figure S8:**
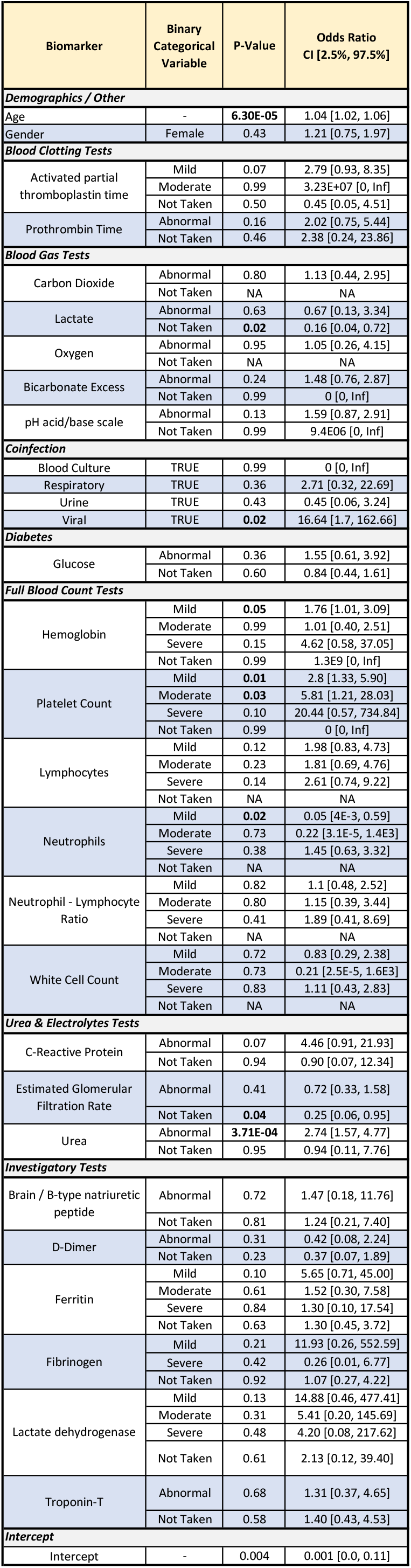
Standard logistic regression odds ratio and confidence intervals per biomarker using all valid biomarker training data available (n=590). Note most biomarkers include a ‘Test Not Taken’ stand in variable.

**Figure S9:**
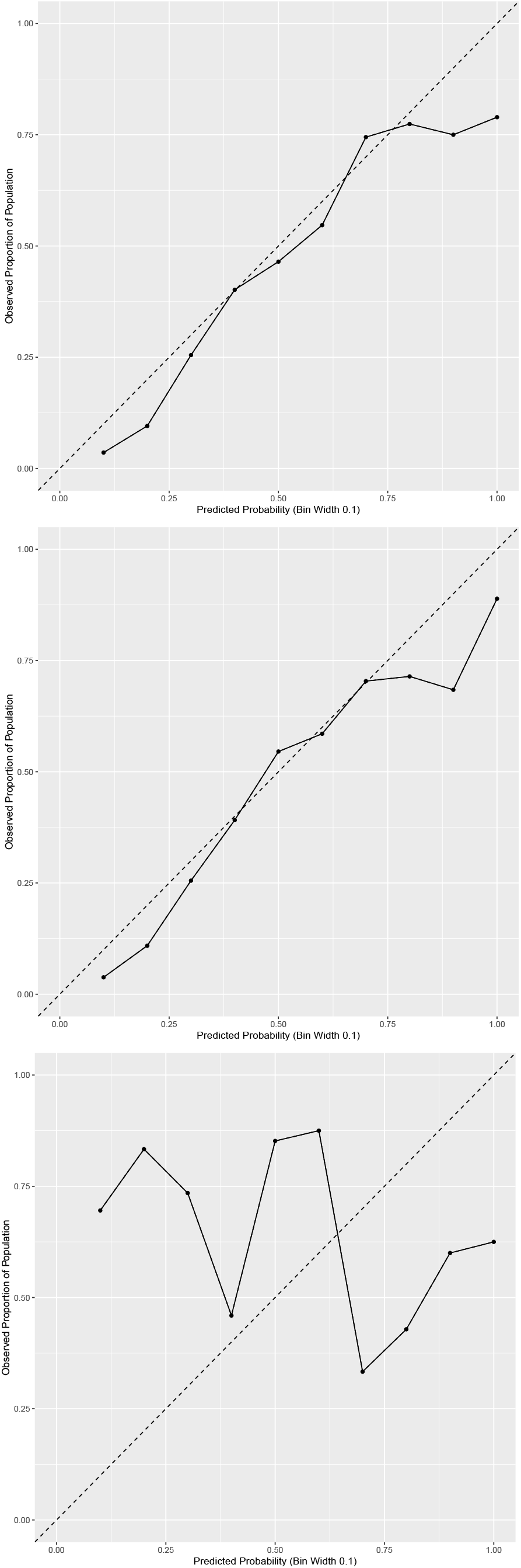
Model calibration depicting a standard GLM model trained on: (Top) all data and tested on all data (Middle); training data (n=590) and tested on the same; (Bottom) training data and tested on validation data (n=293). A well calibrated model should evenly distribute outcome probabilities, i.e. be close to unity.

**Figure S10:**
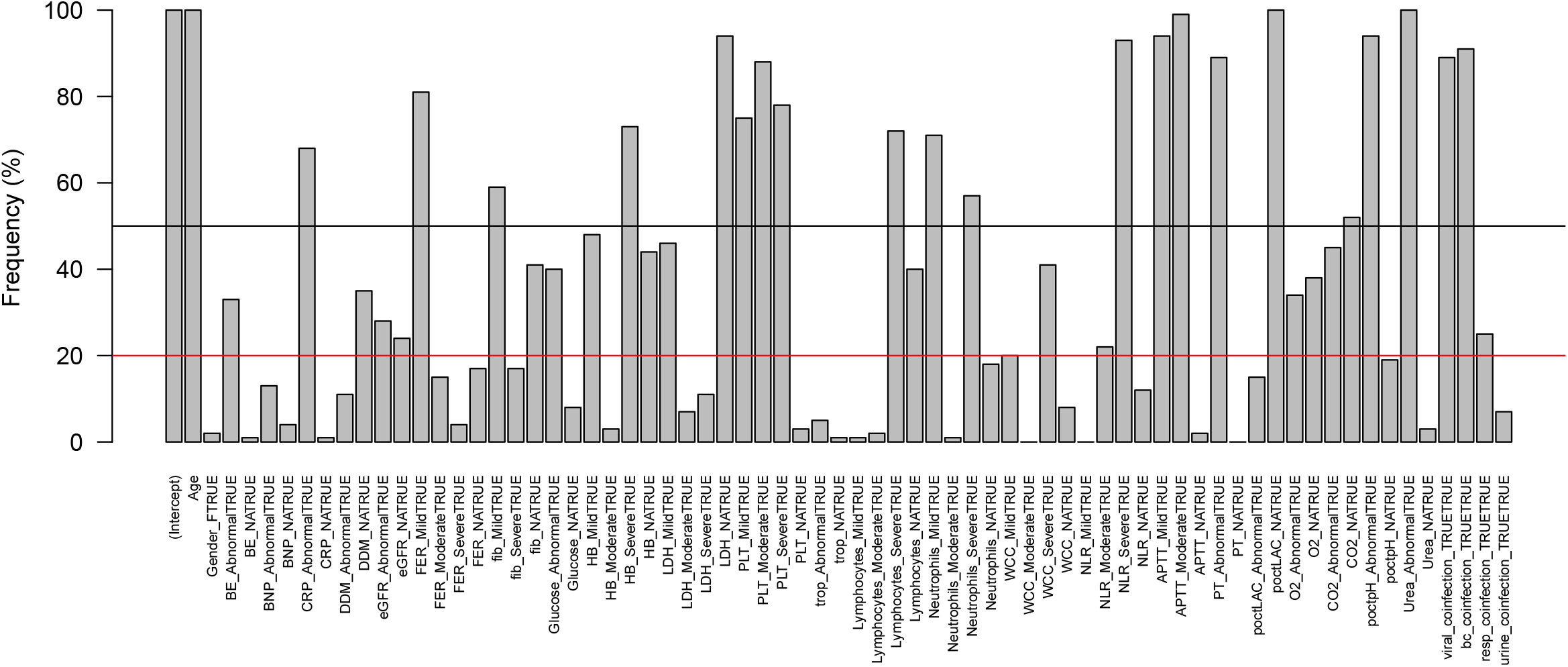
Frequency of LASSO logistic regression variables having a coefficient greater or less than 0. Red and black lines indicate thresholds for 20% and 50% frequency.

**Figure S11:**
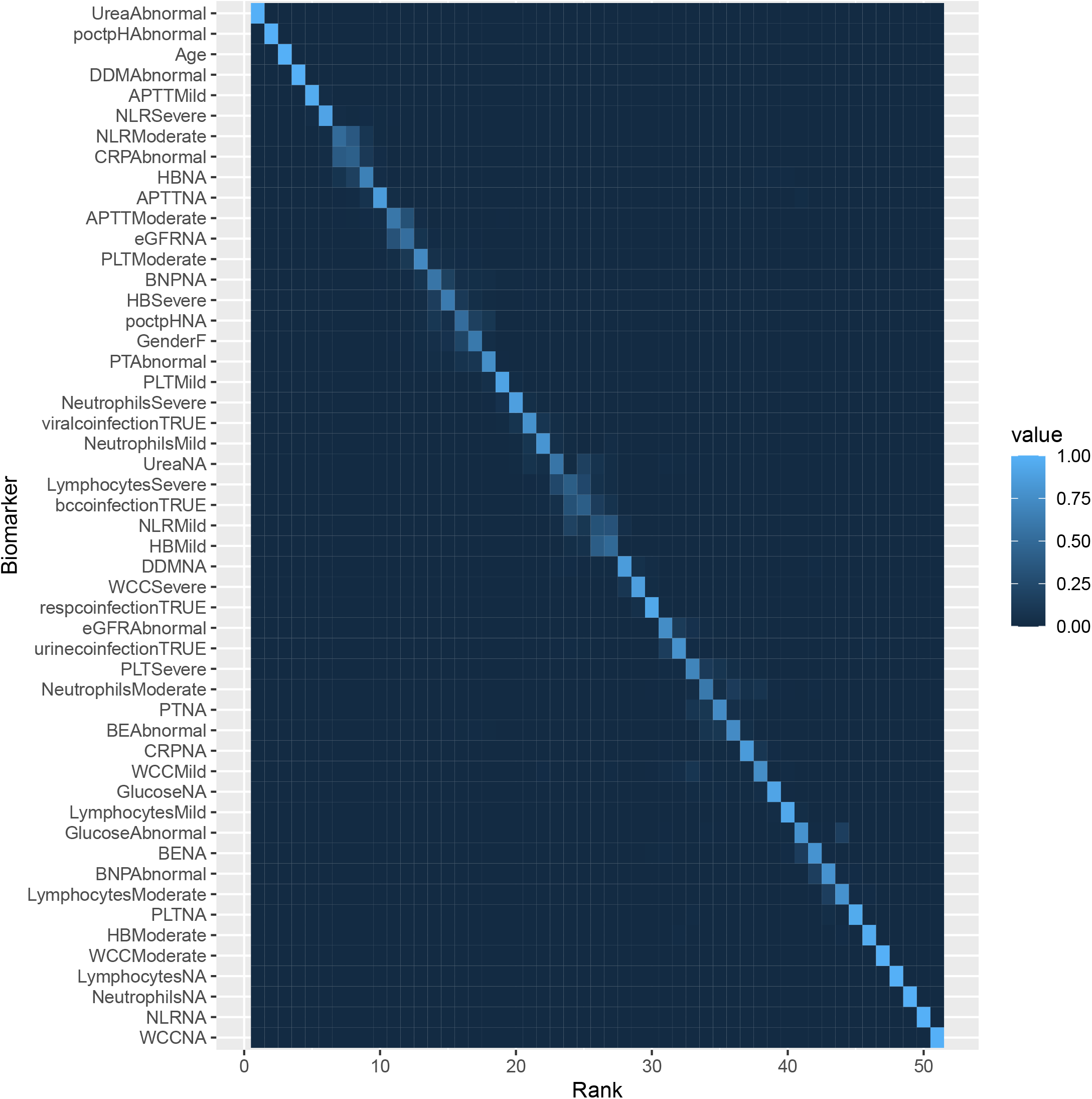
Heatmap representation of the LOO variable selection output from Bayesian projective prediction ranking predictive power as a function in change of AUC. The color of an individual cell shows the proportion of times in the LOO process a variable was chosen at that particular rank of predictive strength. Note this demonstrates a reduced 15-biomarker model (51 variables total), where biomarkers that had ‘Test Not Taken’ ranked as their most important predictive element were removed from the model.

**Figure S12:**
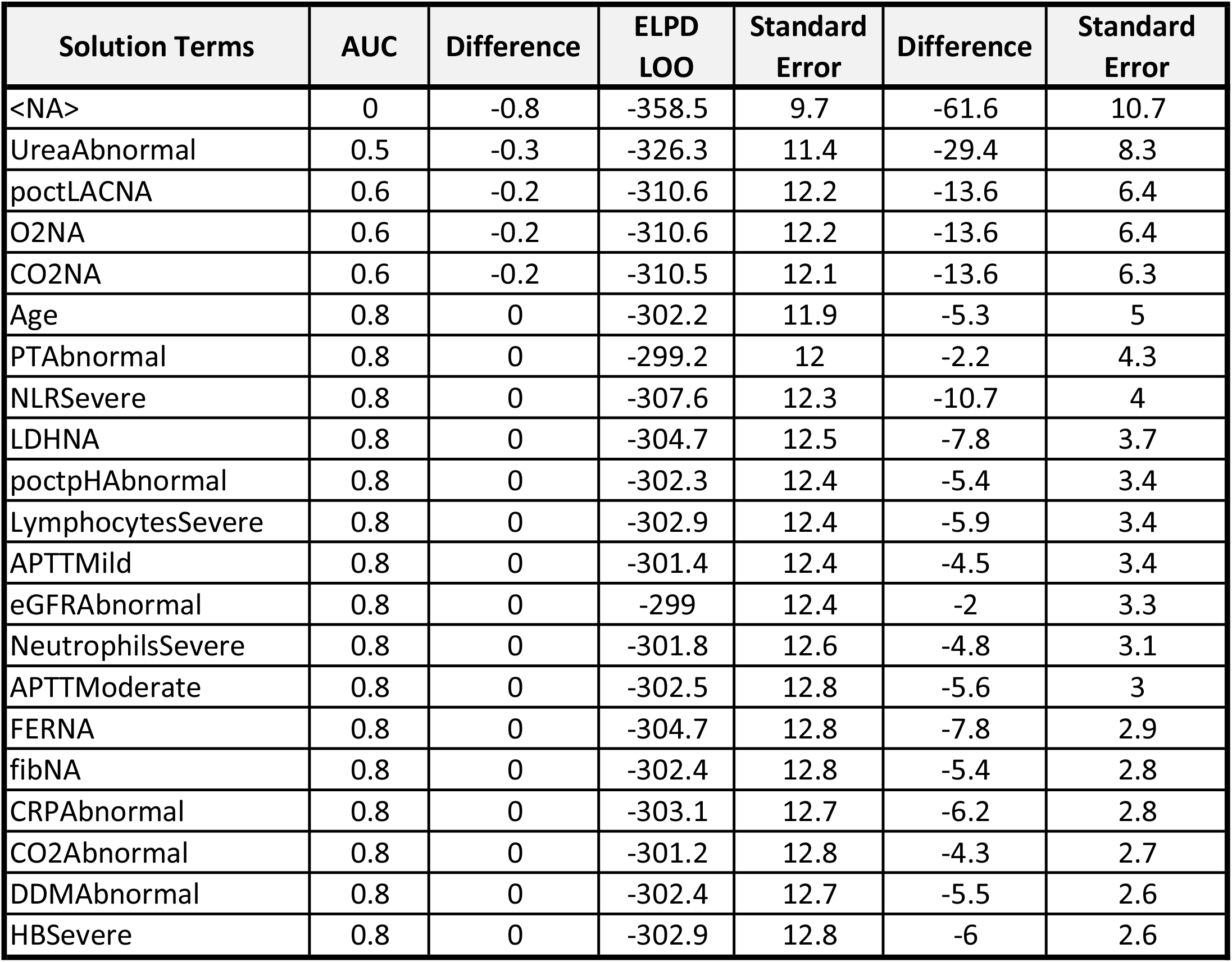
Summary statistics of Bayesian projective prediction ranking the contribution of each variable by change in AUC and expected log-predictive density (ELPD)

**Figure S13:**
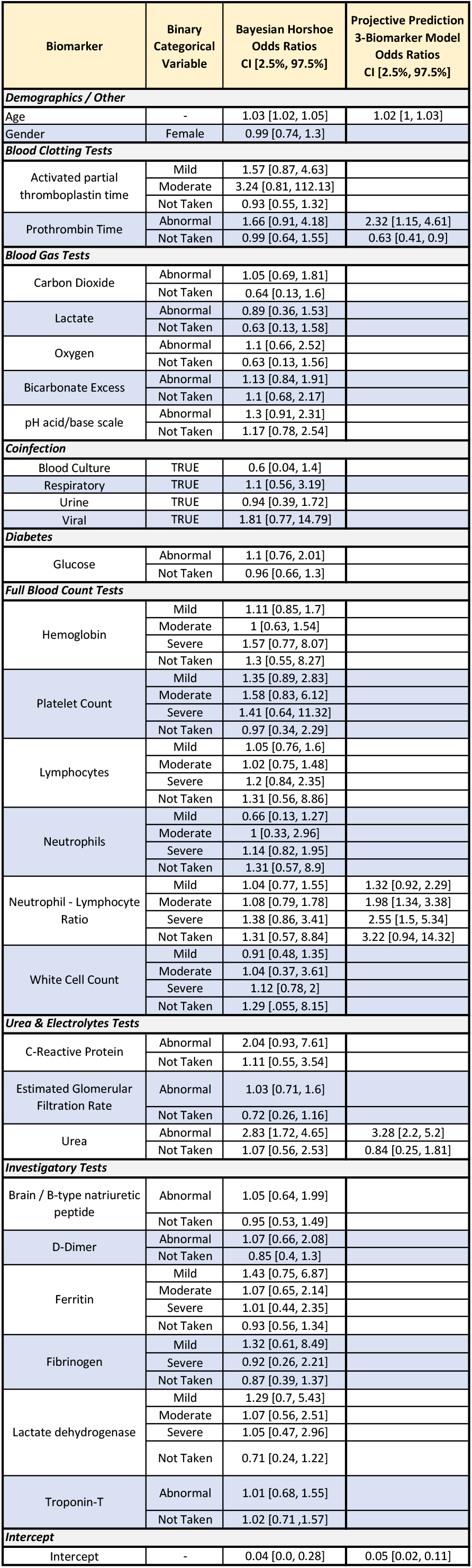
Odds ratios for full Bayesian model and reduced 3-biomarker model via projective prediction

**Figure S14:**
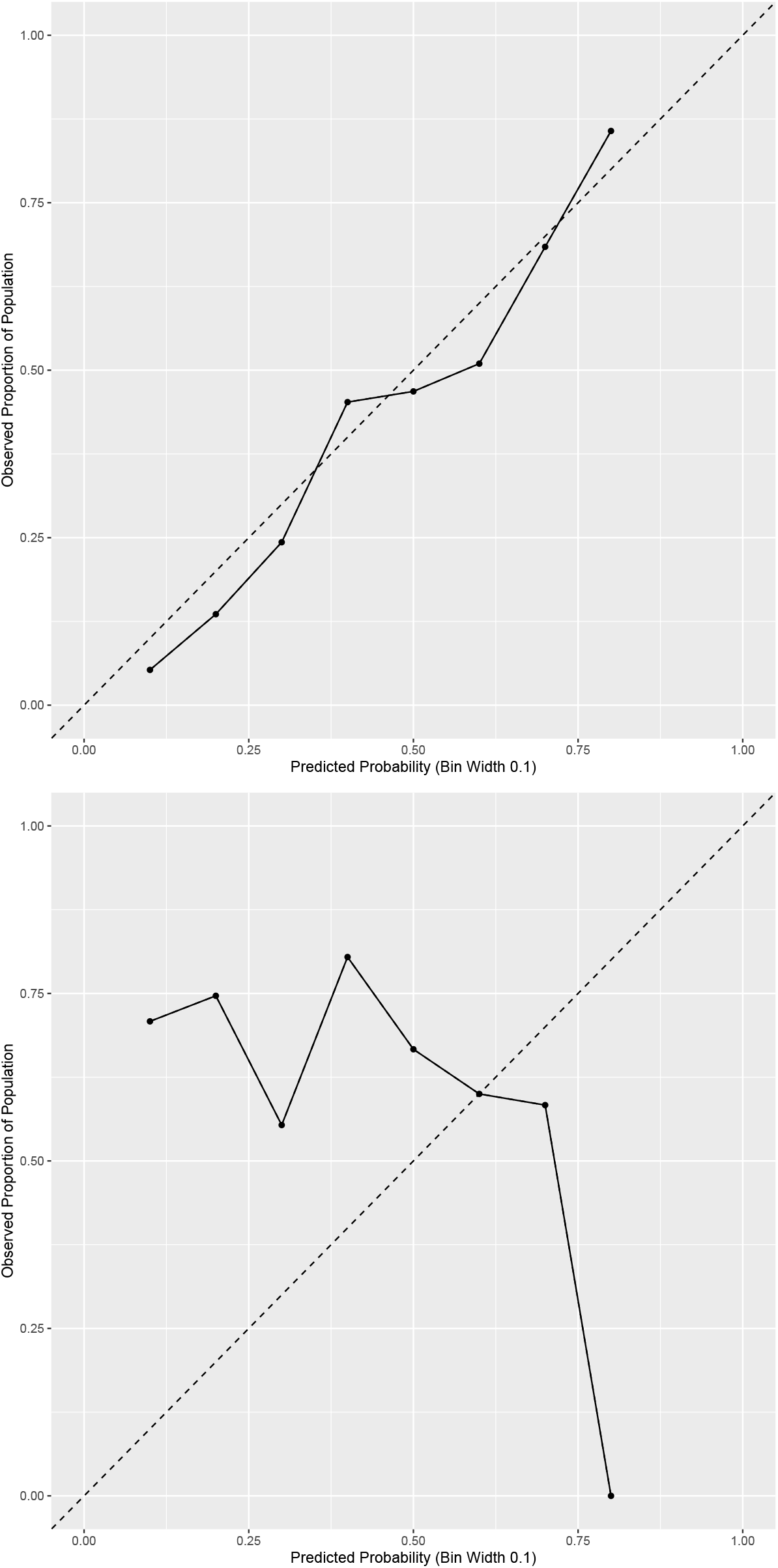
Model calibration depicting Projective Prediction 3-biomarker model tested on: (Top) training data (n=590); (Bottom) validation data (n=293). Note the models do not have points for each of the 10 probability bins because some ranges, e.g. 0.9-1.0 had no patients in this band as judged by the model output. A well calibrated model should evenly distribute outcome probabilities, i.e. be close to unity.

## Notes

### Competing Interest Statement

The authors have declared no competing interest.

### Author Declarations

The study [IRAS project ID: 283439] underwent a rigorous ethical and regulatory approval process, and a favourable opinion was gained from Research Ethics Service, Wales REC 7, c/o Public Health Wales, Building 1, Jobswell Road, St Davids Park, SA31 3HB on 11/09/2020.

### Summary of Updates

Revised title and abstract. Revised cross-validation results and description for stratified approach. Revised cross-validation tables. Added discussion of multiple comparison error

